## Supplemental Information for "Tissue of Origin Characterization of Cell Free DNA in Seminal Plasma: Implications for New Liquid Biopsies"

**Fig. S1.**

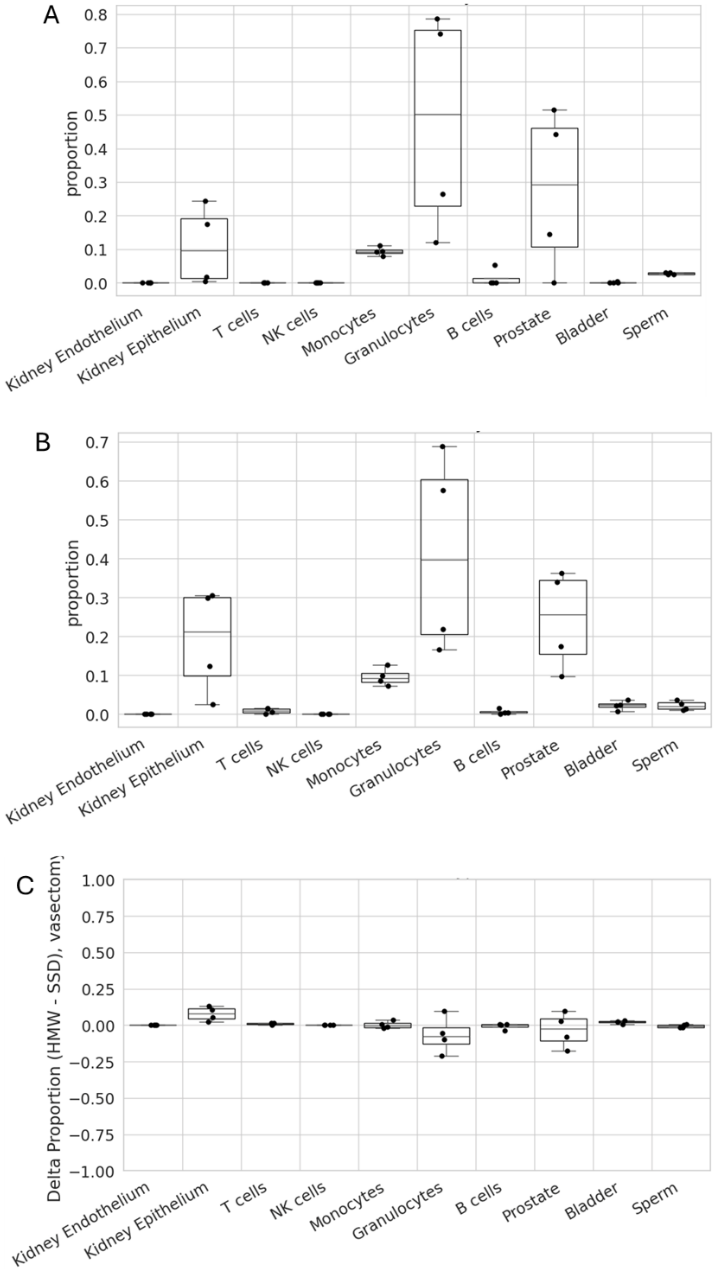


**Tissue Deconvolution Results for Vasectomy Subjects**
A: Tissue Deconvolution Results for SSD
B: Tissue Deconvolution Results for HMW
C: Difference in Proportion of Signal Between HMW and SDD (n=4)

It is important to note that the algorithm used in the tissue deconvolution sums the signal to add to 1 (i.e.100%), therefore while it appears that the vasectomy subjects have a lot more signal from the various somatic cells, this is driven by the absence of any sperm signal.

**Fig. S2**

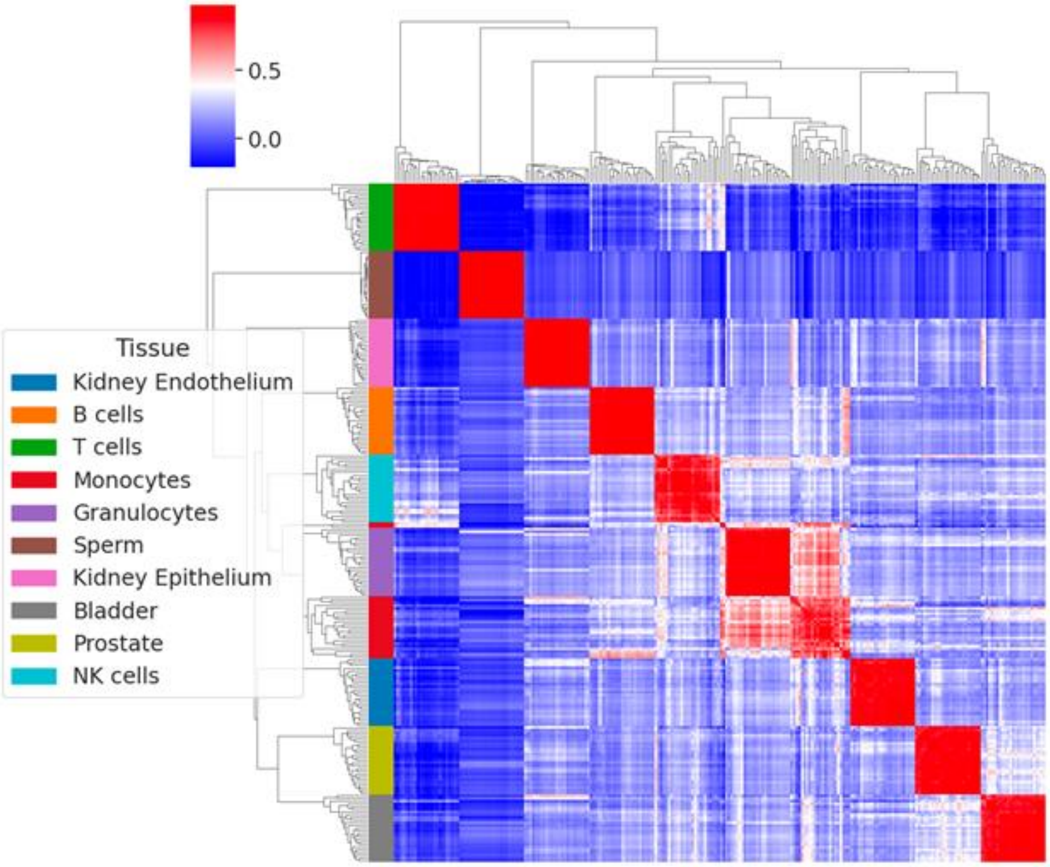


**Unsupervised clustering of reference dataset using our deconvolution signature matrix.** Pearson correlation followed by clustering of the n=25 methylation markers for each reference tissue show strong agreement within their tissue and mostly poor correlation across tissues. An exception is the slight intermixing of the granulocyte/monocyte marker clusters, which can be explained by their similar developmental origins.

**Fig. S3**

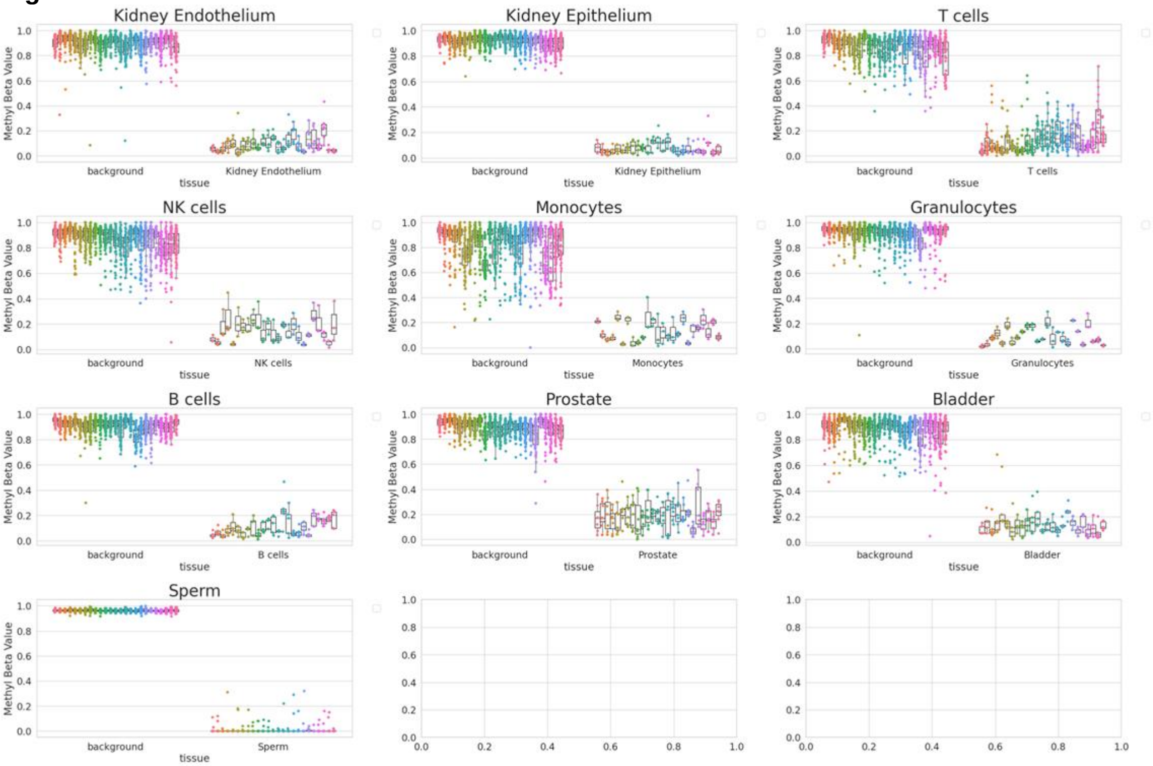


**Methylation values for tissue/cell type specific hypomethylation markers in reference dataset.** Each graph represents methylation values for a tissue/cell type specific set of hypomethylation markers. Shown are methylation values for background (all other tissue/cell types) and the tissue/cell type of interest for all samples in the reference dataset.

**Fig. S4**

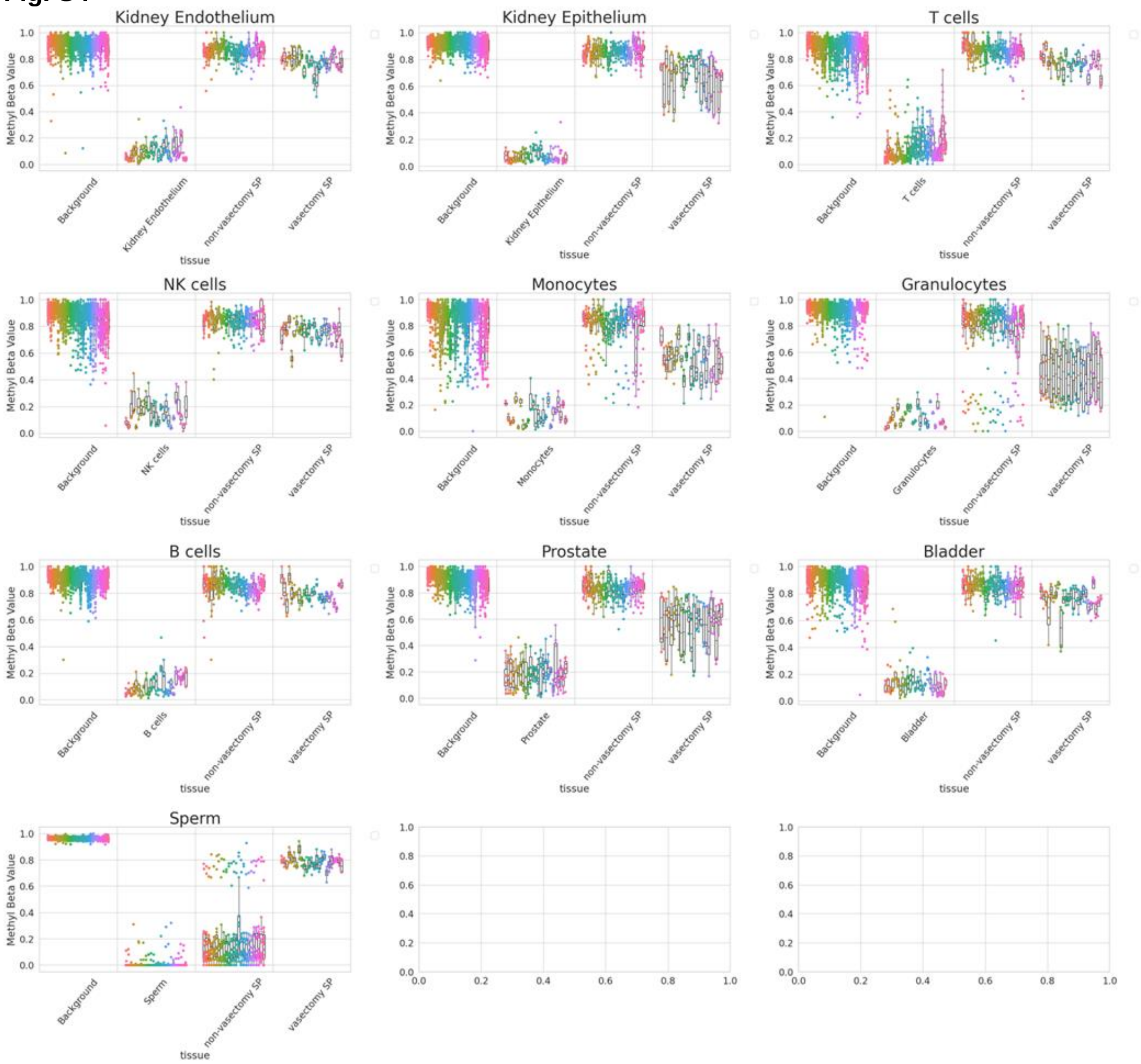


**Methylation values for tissue/cell type specific hypomethylation markers in SSD.** Each graph represents methylation values for a tissue/cell type specific set of hypomethylation markers. Shown are methylation values for background (reference dataset), tissue of interest (reference dataset), non-vasectomy seminal plasma (SP) samples and vasectomy SP samples. Note that the methylation profile for most of the non-vasectomy SP more closely matches sperm more so than any other tissue type, illustrating sperm to be the most prominent cell type. In vasectomy samples, prostate, granulocytes and monocytes markers appear hypothemethylated compared to background tissues, suggesting the presence of DNA from these cell types within these samples.

**Fig. S5**

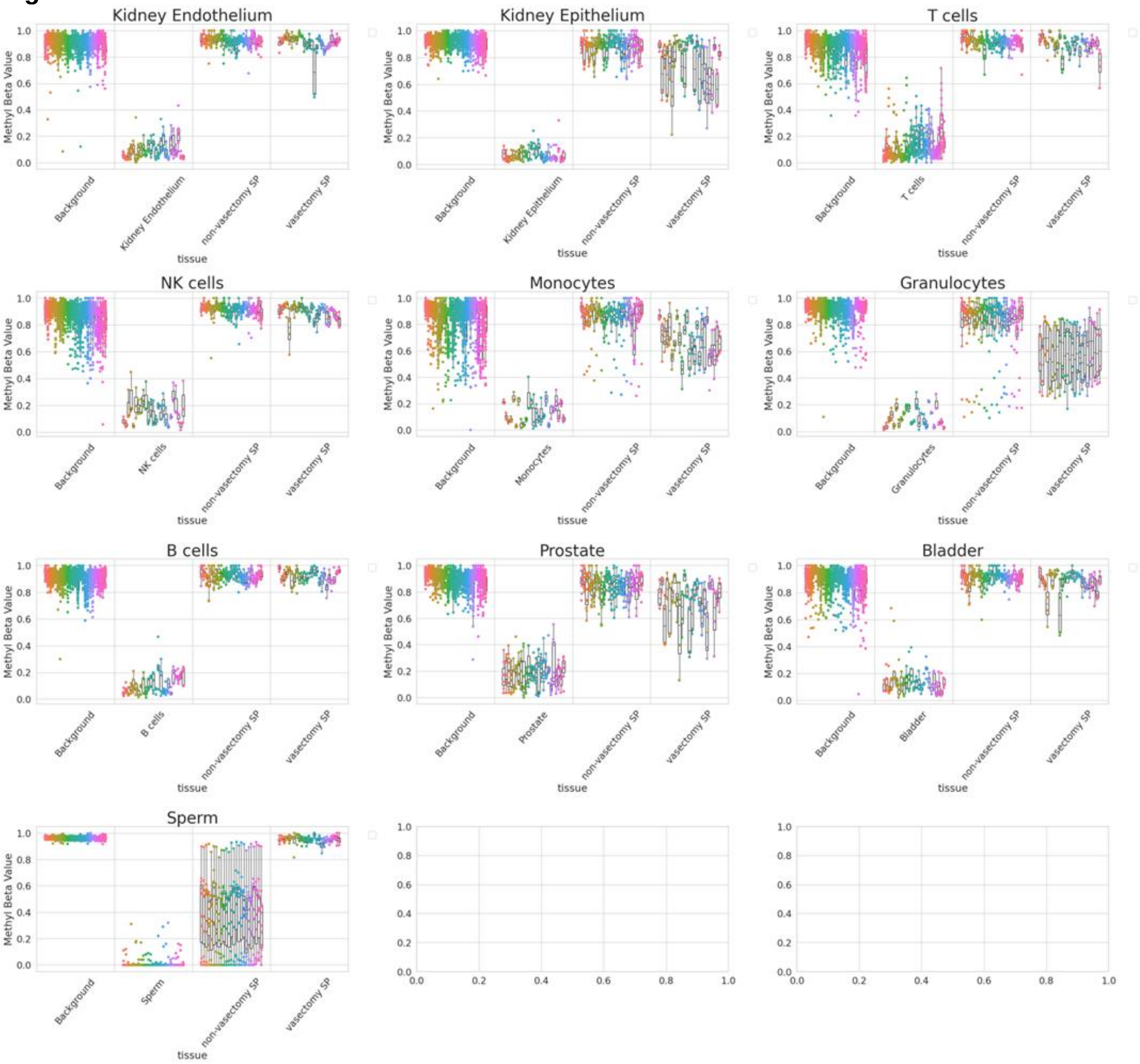


**Methylation values for tissue/cell type specific hypomethylation markers in HMW.** Each graph represents methylation values for a tissue/cell type specific set of hypomethylation markers. Shown are methylation values for background (reference dataset), tissue of interest (reference dataset), non-vasectomy seminal plasma (SP) samples and vasectomy SP samples. Note the very large variability in signal for sperm markers in non-vasectomy samples demonstrating a large range of sperm signal present.

**Table S1: Reference datasets used for tissue deconvolution** (1–4).

| sample_id | tissue | cell_type | source | data_used |
|---|---|---|---|---|
| GSM5652184 | Kidney Endothelium | Kidney glomerular:Endothelium | GSE186458 | preprocessed |
| GSM5652185 | Kidney Endothelium | Kidney glomerular:Endothelium | GSE186458 | preprocessed |
| GSM5652186 | Kidney Endothelium | Kidney glomerular:Endothelium | GSE186458 | preprocessed |
| GSM5652187 | Kidney Endothelium | Kidney tubular:Endothelium | GSE186458 | preprocessed |
| GSM5652188 | Kidney Endothelium | Kidney tubular:Endothelium | GSE186458 | preprocessed |
| GSM5652189 | Kidney Endothelium | Kidney tubular:Endothelium | GSE186458 | preprocessed |
| GSM5652256 | Kidney Epithelium | Kidney glomerular:Epithelium | GSE186458 | preprocessed |
| GSM5652257 | Kidney Epithelium | Kidney glomerular:Epithelium | GSE186458 | preprocessed |
| GSM5652258 | Kidney Epithelium | Kidney tubular:Epithelium | GSE186458 | preprocessed |
| GSM5652259 | Kidney Epithelium | Kidney tubular:Epithelium | GSE186458 | preprocessed |
| GSM5652260 | Kidney Epithelium | Kidney tubular:Epithelium | GSE186458 | preprocessed |
| GSM5652277 | T cells | Blood:T (CD3+) cells | GSE186458 | preprocessed |
| GSM5652278 | T cells | Blood:T (CD3+) cells | GSE186458 | preprocessed |
| GSM5652279 | T cells | Blood:T helper(CD4+) cells | GSE186458 | preprocessed |
| GSM5652280 | T cells | Blood:T helper(CD4+) cells | GSE186458 | preprocessed |
| GSM5652281 | T cells | Blood:T helper(CD4+) cells | GSE186458 | preprocessed |
| GSM5652282 | T cells | Blood:T cytotoxic (CD8+) cells | GSE186458 | preprocessed |
| GSM5652283 | T cells | Blood:T cytotoxic (CD8+) cells | GSE186458 | preprocessed |
| GSM5652284 | T cells | Blood:T cytotoxic (CD8+) cells | GSE186458 | preprocessed |
| GSM5652285 | T cells | Blood:T central memory CD4 | GSE186458 | preprocessed |
| GSM5652286 | T cells | Blood:T central memory CD4 | GSE186458 | preprocessed |
| GSM5652287 | T cells | Blood:T central memory CD4 | GSE186458 | preprocessed |
| GSM5652288 | T cells | Blood:T effector cell CD8 | GSE186458 | preprocessed |
| GSM5652289 | T cells | Blood:T effector cell CD8 | GSE186458 | preprocessed |
| GSM5652290 | T cells | Blood:T effector cell CD8 | GSE186458 | preprocessed |
| GSM5652291 | T cells | Blood:T effector memory CD4 | GSE186458 | preprocessed |
| GSM5652292 | T cells | Blood:T effector memory CD4 | GSE186458 | preprocessed |
| GSM5652293 | T cells | Blood:T effector memory CD4 | GSE186458 | preprocessed |
| GSM5652294 | T cells | Blood:T effector memory CD8 | GSE186458 | preprocessed |
| GSM5652295 | T cells | Blood:T effector memory CD8 | GSE186458 | preprocessed |
| GSM5652296 | T cells | Blood:Naive T cells CD4 | GSE186458 | preprocessed |
| GSM5652297 | T cells | Blood:Naive T cells CD8 | GSE186458 | preprocessed |
| GSM5652298 | T cells | Blood:Naive T cells CD8 | GSE186458 | preprocessed |
| GSM5652299 | NK cells | Blood:NK | GSE186458 | preprocessed |
| GSM5652300 | NK cells | Blood:NK | GSE186458 | preprocessed |
| GSM5652301 | NK cells | Blood:NK | GSE186458 | preprocessed |
| GSM5652302 | Monocytes | Blood:Monocytes | GSE186458 | preprocessed |

| | | | | |
|---|---|---|---|---|
| GSM5652303 | Monocytes | Blood:Monocytes | GSE186458 | preprocessed |
| GSM5652304 | Monocytes | Blood:Monocytes | GSE186458 | preprocessed |
| GSM5652313 | Granulocytes | Blood:Granulocytes | GSE186458 | preprocessed |
| GSM5652314 | Granulocytes | Blood:Granulocytes | GSE186458 | preprocessed |
| GSM5652315 | Granulocytes | Blood:Granulocytes | GSE186458 | preprocessed |
| GSM5652316 | B cells | Blood:B cells | GSE186458 | preprocessed |
| GSM5652317 | B cells | Blood:B cells | GSE186458 | preprocessed |
| GSM5652318 | B cells | Blood:B cells | GSE186458 | preprocessed |
| GSM5652319 | B cells | Blood:Memory B cells | GSE186458 | preprocessed |
| GSM5652320 | B cells | Blood:Memory B cells | GSE186458 | preprocessed |
| GSM5652338 | Prostate | Prostate:Epithelium | GSE186458 | preprocessed |
| GSM5652339 | Prostate | Prostate:Epithelium | GSE186458 | preprocessed |
| GSM5652340 | Prostate | Prostate:Epithelium | GSE186458 | preprocessed |
| GSM5652341 | Prostate | Prostate:Epithelium | GSE186458 | preprocessed |
| GSM5652342 | Bladder | Bladder:Epithelium | GSE186458 | preprocessed |
| GSM5652343 | Bladder | Bladder:Epithelium | GSE186458 | preprocessed |
| GSM5652344 | Bladder | Bladder:Epithelium | GSE186458 | preprocessed |
| GSM5652345 | Bladder | Bladder:Epithelium | GSE186458 | preprocessed |
| GSM5652346 | Bladder | Bladder:Epithelium | GSE186458 | preprocessed |
| SRR6156035 | Prostate | Prostate:Epithelium | PRJNA413837 | raw |
| SRR6156036 | Prostate | Prostate:Epithelium | PRJNA413837 | raw |
| SRR6156037 | Prostate | Prostate:Epithelium | PRJNA413837 | raw |
| SRR10247211 | Sperm | Sperm:Sperm | PRJNA576485 | raw |
| SRR10247197 | Sperm | Sperm:Sperm | PRJNA576485 | raw |
| SRR10247226 | Sperm | Sperm:Sperm | PRJNA576485 | raw |
| SRR10247238 | Sperm | Sperm:Sperm | PRJNA576485 | raw |
| SRR15427078 | Sperm | Sperm:Sperm | PRJNA754049 | raw |
| SRR15427176 | Sperm | Sperm:Sperm | PRJNA754049 | raw |
| SRR15427084 | Sperm | Sperm:Sperm | PRJNA754049 | raw |
| SRR15427086 | Sperm | Sperm:Sperm | PRJNA754049 | raw |
| SRR15427180 | Sperm | Sperm:Sperm | PRJNA754049 | raw |

**Table S2: Signature matrix**

| methyl_region | Kidney Endothelium | Kidney Epithelium | T cells | NK cells | Monocytes | Granulocytes | B cells | Prostate | Bladder | Sperm |
|---|---|---|---|---|---|---|---|---|---|---|
| chr1:1128037-1128257 | 0.85783456 | 0.04317647 | 0.91933088 | 0.92764706 | 0.9519375 | 0.95482353 | 0.938625 | 0.87952941 | 0.88858824 | 0.93 |
| chr1:1221985-1222423 | 0.89584 | 0.912 | 0.87562 | 0.145016 | 0.92084 | 0.93136 | 0.91644 | 0.82 | 0.69416 | 0.94 |
| chr1:1222591-1222811 | 0.9307 | 0.9483 | 0.9365 | 0.12627 | 0.9442 | 0.9266 | 0.9236 | 0.9208 | 0.883 | 0.94 |
| chr1:9022419-9022539 | 0.8949 | 0.605 | 0.9046 | 0.9486 | 0.9692 | 0.952 | 0.9412 | 0.82 | 0.0613 | 0.96 |
| chr1:47993454-47993674 | 0.9148 | 0.06922 | 0.8774 | 0.9182 | 0.9314 | 0.9286 | 0.9414 | 0.9132 | 0.8738 | 0.96 |
| chr1:53438263-53439585 | 0.95693351 | 0.96422727 | 0.95317257 | 0.96261947 | 0.96612963 | 0.96400943 | 0.95946018 | 0.96200901 | 0.95871963 | 0 |
| chr1:54257644-54257764 | 0.10166 | 0.8428 | 0.9011 | 0.912 | 0.967 | 0.9674 | 0.9344 | 0.89 | 0.882 | 0.94 |
| chr1:58928585-58928705 | 0.9218 | 0.916 | 0.935175 | 0.9262 | 0.971 | 0.9518 | 0.92325 | 0.9334 | 0.11292 | 0.94 |
| chr1:90893412-90894018 | 0.95527421 | 0.95475 | 0.96392708 | 0.96504082 | 0.96582979 | 0.96279167 | 0.96970213 | 0.96173469 | 0.95829787 | 0 |
| chr1:116475357-116475568 | 0.90846429 | 0.0271 | 0.9455 | 0.96633333 | 0.94142857 | 0.94614286 | 0.93085714 | 0.87857143 | 0.82128571 | 0.96 |
| chr1:149931235-149931435 | 0.90705 | 0.9697 | 0.93466667 | 0.9351 | 0.20328 | 0.94188889 | 0.94744444 | 0.939 | 0.9256 | 0.95 |
| chr1:159954268-159954787 | 0.72823571 | 0.75342857 | 0.80885714 | 0.81957143 | 0.23934286 | 0.61492857 | 0.78785714 | 0.89 | 0.72185714 | 0.96 |
| chr1:164682675-164682891 | 0.908 | 0.09408 | 0.9334 | 0.9262 | 0.9618 | 0.9452 | 0.9336 | 0.9302 | 0.954 | 0.96 |
| chr1:167485529-167485749 | 0.7344 | 0.84 | 0.7718 | 0.15224 | 0.9198 | 0.9206 | 0.813 | 0.8054 | 0.7358 | 0.9 |
| chr1:209838583-209838803 | 0.8821 | 0.8496 | 0.9474 | 0.957 | 0.9368 | 0.08198 | 0.9566 | 0.9 | 0.9378 | 0.97 |
| chr1:219613125-219613245 | 0.97720833 | 0.9777 | 0.96875 | 0.96488889 | 0.979125 | 0.9557 | 0.97622222 | 0.97 | 0.96477778 | 0 |
| chr1:232590982-232591202 | 0.4404 | 0.9506 | 0.8141 | 0.9334 | 0.0829 | 0.5134 | 0.73 | 0.9476 | 0.9532 | 0.95 |
| chr1:244941212-244941332 | 0.96841667 | 0.96733333 | 0.95266667 | 0.95283333 | 0.94616667 | 0.97366667 | 0.04961667 | 0.96 | 0.9705 | 0.96 |
| chr2:3389191-3389659 | 0.95781471 | 0.95997143 | 0.96317143 | 0.96782857 | 0.97231429 | 0.97237143 | 0.96522857 | 0.9634 | 0.96485714 | 0 |

| | | | | | | | | | | |
|---|---|---|---|---|---|---|---|---|---|---|
| chr2:3533318-3533622 | 0.92241667 | 0.947 | 0.92841667 | 0.917 | 0.9705 | 0.96216667 | 0.937 | 0.24 | 0.9025 | 0.91 |
| chr2:8282949-8283069 | 0.9381 | 0.9232 | 0.6965 | 0.0345 | 0.9574 | 0.9558 | 0.9182 | 0.87 | 0.8952 | 0.94 |
| chr2:12767709-12767895 | 0.89291667 | 0.90666667 | 0.931 | 0.96233333 | 0.958 | 0.92916667 | 0.93466667 | 0.17666667 | 0.824 | 0.95 |
| chr2:29224599-29224719 | 0.89558333 | 0.91583333 | 0.909 | 0.94883333 | 0.92616667 | 0.937 | 0.904 | 0.86 | 0.18298333 | 0.92 |
| chr2:105881290-105882163 | 0.96545385 | 0.96595062 | 0.96247608 | 0.97175641 | 0.96853165 | 0.97183951 | 0.96353333 | 0.97 | 0.96375 | 0 |
| chr2:109700376-109700496 | 0.06483 | 0.9378 | 0.8566 | 0.8824 | 0.907 | 0.9192 | 0.8446 | 0.94 | 0.879 | 0.97 |
| chr2:128436859-128436979 | 0.906 | 0.9056 | 0.07974 | 0.4654 | 0.9602 | 0.948 | 0.8564 | 0.9226 | 0.9088 | 0.92 |
| chr2:231528339-231528559 | 0.94214286 | 0.90064286 | 0.93462637 | 0.07200714 | 0.94784615 | 0.92014286 | 0.91914286 | 0.89742857 | 0.88561538 | 0.96 |
| chr2:233761726-233761983 | 0.8917 | 0.9056 | 0.8565 | 0.8754 | 0.9362 | 0.9084 | 0.8808 | 0.79 | 0.08166 | 0.96 |
| chr2:241925322-241925542 | 0.83379167 | 0.91408333 | 0.902625 | 0.205325 | 0.94981818 | 0.9475 | 0.93925 | 0.92016667 | 0.94208333 | 0.94 |
| chr3:4815199-4815490 | 0.942 | 0.9589 | 0.93285 | 0.23033 | 0.9195 | 0.9038 | 0.9557 | 0.9539 | 0.9351 | 0.93 |
| chr3:15451108-15451328 | 0.893 | 0.897 | 0.89035714 | 0.04724286 | 0.91357143 | 0.915 | 0.91442857 | 0.94 | 0.93128571 | 0.95 |
| chr3:49683289-49683495 | 0.8575 | 0.04772 | 0.9577 | 0.9732 | 0.9554 | 0.9556 | 0.9518 | 0.8228 | 0.9194 | 0.94 |
| chr3:119311066-119311286 | 0.8705 | 0.9265 | 0.92608333 | 0.9295 | 0.54966667 | 0.07275 | 0.91416667 | 0.942 | 0.90033333 | 0.94 |
| chr3:129590404-129590579 | 0.09011667 | 0.873 | 0.93841667 | 0.9425 | 0.79933333 | 0.872 | 0.94933333 | 0.91733333 | 0.93133333 | 0.93 |
| chr3:130919068-130919188 | 0.9695 | 0.972 | 0.961 | 0.9662 | 0.9616 | 0.9866 | 0.9764 | 0.96 | 0.9656 | 0 |
| chr3:153067100-153067265 | 0.93055556 | 0.94855556 | 0.88022222 | 0.91211111 | 0.61675 | 0.05873333 | 0.90175 | 0.95144444 | 0.94877778 | 0.97 |
| chr3:169931772-169931917 | 0.9144 | 0.8646 | 0.9069 | 0.9132 | 0.9594 | 0.9588 | 0.09058 | 0.9136 | 0.9094 | 0.94 |
| chr3:186996123-186996667 | 0.93866667 | 0.9375 | 0.92575 | 0.92133333 | 0.91666667 | 0.90316667 | 0.19698333 | 0.96016667 | 0.94816667 | 0.94 |

| | | | | | | | | | | |
|---|---|---|---|---|---|---|---|---|---|---|
| chr4:2959628-2959826 | 0.91283333 | 0.95266667 | 0.93383333 | 0.94083333 | 0.9695 | 0.95316667 | 0.97233333 | 0.93 | 0.12578333 | 0.93 |
| chr4:42361556-42361776 | 0.91378571 | 0.05354286 | 0.88407143 | 0.91985714 | 0.93085714 | 0.93314286 | 0.92314286 | 0.93 | 0.92442857 | 0.92 |
| chr4:151701648-151701918 | 0.945 | 0.96633333 | 0.95216667 | 0.97116667 | 0.74533333 | 0.17295 | 0.96766667 | 0.96 | 0.96516667 | 0.96 |
| chr4:153022212-153022432 | 0.92027778 | 0.939 | 0.91955556 | 0.91811111 | 0.95533333 | 0.92944444 | 0.96588889 | 0.17 | 0.853 | 0.96 |
| chr4:153659402-153659522 | 0.9619 | 0.8598 | 0.9069 | 0.9436 | 0.23642 | 0.4872 | 0.9408 | 0.94 | 0.9484 | 0.97 |
| chr5:1282490-1282669 | 0.81309091 | 0.89263636 | 0.90072727 | 0.90936364 | 0.90263636 | 0.92754545 | 0.93027273 | 0.19479091 | 0.79109091 | 0.93 |
| chr5:1564297-1564417 | 0.745 | 0.72166667 | 0.73566667 | 0.6005 | 0.02185 | 0.49856667 | 0.63583333 | 0.71 | 0.60983333 | 0.93 |
| chr5:14325814-14326122 | 0.9291 | 0.9102 | 0.8581 | 0.9164 | 0.8468 | 0.03978 | 0.938 | 0.947 | 0.9462 | 0.94 |
| chr5:32098360-32099276 | 0.09977353 | 0.81764706 | 0.89344118 | 0.92629412 | 0.87947059 | 0.87735294 | 0.89835294 | 0.85447059 | 0.91670588 | 0.93 |
| chr5:72301763-72301983 | 0.9236 | 0.8778 | 0.9531 | 0.9538 | 0.9808 | 0.9576 | 0.20522 | 0.9368 | 0.941 | 0.92 |
| chr5:73508612-73508818 | 0.96065217 | 0.95856522 | 0.96498701 | 0.96717391 | 0.96659091 | 0.96833333 | 0.96716667 | 0.96 | 0.969 | 0 |
| chr5:99410507-99410676 | 0.95595588 | 0.96905882 | 0.96042647 | 0.9506 | 0.96822222 | 0.9679375 | 0.9675625 | 0.96 | 0.96705556 | 0 |
| chr5:135127721-135128159 | 0.91745455 | 0.80145455 | 0.87822727 | 0.95654545 | 0.93827273 | 0.94172727 | 0.91927273 | 0.71 | 0.11567273 | 0.94 |
| chr5:139740420-139740640 | 0.73796875 | 0.860875 | 0.87746875 | 0.7196875 | 0.900625 | 0.89375 | 0.065225 | 0.91 | 0.852375 | 0.89 |
| chr5:172156734-172156944 | 0.0733875 | 0.819 | 0.9539375 | 0.89725 | 0.95075 | 0.959625 | 0.947625 | 0.93525 | 0.9335 | 0.96 |
| chr5:174617359-174618107 | 0.74154167 | 0.86608333 | 0.76370833 | 0.7965 | 0.88066667 | 0.88866667 | 0.81491667 | 0.78 | 0.09605 | 0.94 |
| chr5:174618178-174618321 | 0.7336 | 0.8826 | 0.89 | 0.933 | 0.9286 | 0.9458 | 0.8898 | 0.93 | 0.07974 | 0.95 |
| chr5:178305248-178305404 | 0.89966667 | 0.90166667 | 0.80858333 | 0.90883333 | 0.96433333 | 0.9515 | 0.84533333 | 0.21 | 0.89133333 | 0.94 |
| chr6:1635258-1635697 | 0.06274667 | 0.87213333 | 0.89783333 | 0.91733333 | 0.84093333 | 0.80708 | 0.948 | 0.89 | 0.91766667 | 0.95 |
| chr6:1840809-1840993 | 0.91958333 | 0.94716667 | 0.92808333 | 0.9515 | 0.94833333 | 0.78683333 | 0.94666667 | 0.888 | 0.16835 | 0.89 |
| chr6:15505676-15505896 | 0.91730449 | 0.90408333 | 0.90373077 | 0.03856923 | 0.65930769 | 0.59053846 | 0.91292308 | 0.88638462 | 0.95008333 | 0.95 |

| | | | | | | | | | | |
|---|---|---|---|---|---|---|---|---|---|---|
| chr6:26545718-26545938 | 0.8957 | 0.9516 | 0.9408 | 0.9066 | 0.08992 | 0.6476 | 0.8242 | 0.9166 | 0.9398 | 0.93 |
| chr6:30329300-30329868 | 0.94358824 | 0.95017647 | 0.95432353 | 0.96958824 | 0.84194118 | 0.08642353 | 0.962 | 0.95135294 | 0.95476471 | 0.94 |
| chr6:100829435-100829555 | 0.9569 | 0.9591 | 0.96194444 | 0.9775 | 0.97088889 | 0.9688 | 0.9496 | 0.9617 | 0.9563 | 0 |
| chr6:110705566-110705731 | 0.9518 | 0.9548 | 0.9362 | 0.941 | 0.1756 | 0.899 | 0.4788 | 0.9736 | 0.9764 | 0.91 |
| chr6:114246669-114246875 | 0.9393 | 0.9326 | 0.8895 | 0.898 | 0.9198 | 0.9116 | 0.923 | 0.18 | 0.882 | 0.96 |
| chr6:131575597-131576398 | 0.87457143 | 0.91614286 | 0.93535714 | 0.94642857 | 0.726 | 0.1323 | 0.95314286 | 0.846 | 0.762 | 0.94 |
| chr6:165676764-165676884 | 0.13775 | 0.895 | 0.93591667 | 0.95633333 | 0.93933333 | 0.94816667 | 0.9545 | 0.90666667 | 0.82683333 | 0.96 |
| chr6:170444115-170444335 | 0.83557143 | 0.833 | 0.70623571 | 0.12842857 | 0.76525714 | 0.76014286 | 0.70428571 | 0.76704286 | 0.76432857 | 0.93 |
| chr7:540956-541176 | 0.9155 | 0.952 | 0.7999 | 0.8284 | 0.9154 | 0.9506 | 0.07886 | 0.9486 | 0.9724 | 0.94 |
| chr7:697059-697226 | 0.09277143 | 0.90028571 | 0.90528571 | 0.925 | 0.93742857 | 0.94471429 | 0.94042857 | 0.86 | 0.87357143 | 0.95 |
| chr7:1000212-1000697 | 0.89163462 | 0.86184615 | 0.92211538 | 0.91130769 | 0.89784615 | 0.91607692 | 0.17124231 | 0.87 | 0.91311538 | 0.94 |
| chr7:1004050-1004270 | 0.898 | 0.8632 | 0.93355 | 0.8602 | 0.8657 | 0.897 | 0.0301 | 0.8536 | 0.8913 | 0.91 |
| chr7:1041976-1042115 | 0.83505 | 0.656 | 0.8377 | 0.07716 | 0.87944444 | 0.8876 | 0.92844444 | 0.92 | 0.9327 | 0.93 |
| chr7:1824623-1824743 | 0.87575 | 0.70266667 | 0.09846667 | 0.78133333 | 0.93433333 | 0.93383333 | 0.89233333 | 0.76816667 | 0.77616667 | 0.93 |
| chr7:1824795-1824915 | 0.93473611 | 0.91688889 | 0.13628125 | 0.71755556 | 0.96644444 | 0.94988889 | 0.93277778 | 0.922 | 0.94922222 | 0.96 |
| chr7:1944206-1944426 | 0.72400769 | 0.79009231 | 0.72263077 | 0.19549231 | 0.86461538 | 0.83269231 | 0.86923077 | 0.88 | 0.88730769 | 0.89 |
| chr7:2432468-2434041 | 0.95305823 | 0.95832308 | 0.9627262 | 0.96509023 | 0.96426515 | 0.96918898 | 0.9671746 | 0.96 | 0.962088 | 0 |
| chr7:3107348-3107468 | 0.78141667 | 0.79333333 | 0.79666667 | 0.06231667 | 0.908 | 0.9216 | 0.87383333 | 0.87 | 0.81216667 | 0.86 |
| chr7:3943036-3943256 | 0.91616667 | 0.04368333 | 0.92633333 | 0.94383333 | 0.96133333 | 0.95233333 | 0.9215 | 0.884 | 0.9375 | 0.9 |
| chr7:6622441-6623241 | 0.9537087 | 0.95787805 | 0.96564005 | 0.96736585 | 0.96712195 | 0.97088889 | 0.96456098 | 0.95506173 | 0.95561538 | 0 |
| chr7:6652314-6653106 | 0.95474403 | 0.95698058 | 0.96504248 | 0.97157 | 0.96941748 | 0.96817476 | 0.97107 | 0.96 | 0.95303922 | 0 |
| chr7:47315239-47315459 | 0.04543068 | 0.76175 | 0.86385833 | 0.87525 | 0.89063636 | 0.89641667 | 0.89063636 | 0.80583333 | 0.90108333 | 0.94 |

| | | | | | | | | | | |
|---|---|---|---|---|---|---|---|---|---|---|
| chr7:70416678-70416798 | 0.9497 | 0.93 | 0.9537 | 0.9516 | 0.962 | 0.9728 | 0.234 | 0.96 | 0.9182 | 0.97 |
| chr7:139683163-139683283 | 0.89771429 | 0.0524 | 0.91578571 | 0.94528571 | 0.91314286 | 0.85414286 | 0.80685714 | 0.86385714 | 0.93714286 | 0.96 |
| chr7:151454423-151454543 | 0.9222 | 0.12064 | 0.89465 | 0.9282 | 0.92975 | 0.9312 | 0.8618 | 0.958 | 0.9318 | 0.93 |
| chr7:157630386-157630606 | 0.85368182 | 0.78036364 | 0.89063636 | 0.90463636 | 0.89136364 | 0.88427273 | 0.86809091 | 0.14180909 | 0.687 | 0.93 |
| chr8:11492706-11494456 | 0.72706522 | 0.80330435 | 0.82243478 | 0.8283913 | 0.77951739 | 0.87443478 | 0.05103913 | 0.87213043 | 0.80252174 | 0.92 |
| chr8:22224443-22224663 | 0.77558333 | 0.79883333 | 0.91066667 | 0.94116667 | 0.88216667 | 0.835 | 0.9205 | 0.16216667 | 0.84583333 | 0.97 |
| chr8:23250338-23250558 | 0.7768 | 0.8908 | 0.22766 | 0.756 | 0.8736 | 0.8094 | 0.8386 | 0.8472 | 0.8656 | 0.94 |
| chr8:29061291-29061718 | 0.90822222 | 0.92372222 | 0.15197778 | 0.80894444 | 0.92761111 | 0.95072222 | 0.84316667 | 0.90155556 | 0.8965 | 0.95 |
| chr8:29103709-29103934 | 0.9268125 | 0.939875 | 0.924375 | 0.86975 | 0.074625 | 0.567875 | 0.674875 | 0.9465 | 0.923875 | 0.94 |
| chr8:42437102-42437593 | 0.95406944 | 0.95813889 | 0.96245833 | 0.96874286 | 0.97257143 | 0.96868571 | 0.96833333 | 0.96148571 | 0.9665 | 0 |
| chr8:58070023-58070422 | 0.794 | 0.05229375 | 0.9423125 | 0.93175 | 0.95875 | 0.9530625 | 0.9369375 | 0.951 | 0.946625 | 0.95 |
| chr8:90606379-90606499 | 0.9569 | 0.96741667 | 0.96724176 | 0.96792308 | 0.97692857 | 0.97046667 | 0.97728571 | 0.96 | 0.96185714 | 0 |
| chr8:141152772-141152892 | 0.9213 | 0.9484 | 0.8688 | 0.11162 | 0.6242 | 0.7454 | 0.823 | 0.9506 | 0.8796 | 0.95 |
| chr8:143461298-143462155 | 0.81502931 | 0.83768966 | 0.12584483 | 0.75803571 | 0.91206897 | 0.91882759 | 0.78196552 | 0.79548276 | 0.78496552 | 0.92 |
| chr8:143958802-143959676 | 0.83756008 | 0.81265714 | 0.9046037 | 0.92029412 | 0.92632353 | 0.86848571 | 0.91732353 | 0.24993714 | 0.81768571 | 0.92 |
| chr9:4155306-4155548 | 0.88271429 | 0.04482857 | 0.85021429 | 0.93 | 0.93457143 | 0.94871429 | 0.86642857 | 0.88 | 0.86671429 | 0.92 |
| chr9:33318727-33319100 | 0.94715 | 0.9644 | 0.95755 | 0.9676 | 0.5752 | 0.03418 | 0.9708 | 0.95 | 0.9705 | 0.94 |
| chr9:37324296-37324416 | 0.9017 | 0.9542 | 0.9588 | 0.911 | 0.9576 | 0.9514 | 0.09952 | 0.9418 | 0.9526 | 0.93 |
| chr9:89471485-89472145 | 0.95833333 | 0.96283333 | 0.172025 | 0.76216667 | 0.95366667 | 0.94033333 | 0.73783333 | 0.84066667 | 0.80766667 | 0.94 |
| chr9:92837361-92837481 | 0.9736875 | 0.96525 | 0.9689375 | 0.973625 | 0.970875 | 0.970125 | 0.974375 | 0.968 | 0.980125 | 0 |
| chr9:99998980-99999159 | 0.967575 | 0.96945 | 0.97176316 | 0.97889474 | 0.9745 | 0.97605 | 0.96805263 | 0.96 | 0.9638 | 0 |
| chr9:123608227-123608347 | 0.9211 | 0.9226 | 0.9269 | 0.9224 | 0.8652 | 0.8154 | 0.08266 | 0.93 | 0.9176 | 0.91 |

| | | | | | | | | | | |
|---|---|---|---|---|---|---|---|---|---|---|
| chr9:126885983-126886103 | 0.8969 | 0.7958 | 0.9081 | 0.8162 | 0.6963 | 0.06668 | 0.9294 | 0.93 | 0.8936 | 0.93 |
| chr9:127852062-127852267 | 0.03030714 | 0.68657143 | 0.92278571 | 0.846 | 0.86814286 | 0.85685714 | 0.87985714 | 0.90042857 | 0.93442857 | 0.94 |
| chr9:130118947-130119167 | 0.880875 | 0.0924375 | 0.959875 | 0.957375 | 0.971125 | 0.95225 | 0.940375 | 0.90325 | 0.919625 | 0.93 |
| chr9:132888170-132888390 | 0.934 | 0.95175 | 0.907 | 0.917125 | 0.96125 | 0.943625 | 0.14125 | 0.94425 | 0.920625 | 0.96 |
| chr9:133822414-133822534 | 0.94166667 | 0.94016667 | 0.92041667 | 0.92733333 | 0.95316667 | 0.97466667 | 0.17866667 | 0.94866667 | 0.9235 | 0.93 |
| chr9:136917785-136918015 | 0.89419231 | 0.92630769 | 0.93092308 | 0.95069231 | 0.63792308 | 0.07723077 | 0.92461538 | 0.89 | 0.92192308 | 0.95 |
| chr10:87620-87740 | 0.9136 | 0.947 | 0.9268 | 0.8792 | 0.8912 | 0.8838 | 0.9244 | 0.91 | 0.1441 | 0.96 |
| chr10:1042606-1042809 | 0.92825 | 0.949 | 0.96091667 | 0.94883333 | 0.85883333 | 0.12453333 | 0.95016667 | 0.9525 | 0.9395 | 0.95 |
| chr10:3532772-3532892 | 0.9431 | 0.861 | 0.9001 | 0.9302 | 0.7974 | 0.935 | 0.8634 | 0.91 | 0.159 | 0.95 |
| chr10:6023812-6023932 | 0.9265 | 0.9526 | 0.9565 | 0.954 | 0.9644 | 0.9482 | 0.9628 | 0.17 | 0.8864 | 0.95 |
| chr10:7334089-7334309 | 0.206625 | 0.90475 | 0.9469375 | 0.947125 | 0.955875 | 0.923875 | 0.955 | 0.954625 | 0.93325 | 0.93 |
| chr10:8013792-8013912 | 0.9464 | 0.8284 | 0.9681 | 0.978 | 0.9344 | 0.9692 | 0.956 | 0.9682 | 0.14906 | 0.95 |
| chr10:15037600-15038020 | 0.88233333 | 0.84233333 | 0.8765 | 0.92566667 | 0.9465 | 0.92633333 | 0.85583333 | 0.19 | 0.83166667 | 0.88 |
| chr10:77872230-77872450 | 0.7691 | 0.73332 | 0.8598 | 0.937 | 0.8678 | 0.843 | 0.7758 | 0.061 | 0.7344 | 0.93 |
| chr10:89235430-89235599 | 0.90925 | 0.93533333 | 0.87183333 | 0.9165 | 0.061 | 0.37333333 | 0.939 | 0.94216667 | 0.94366667 | 0.94 |
| chr10:104033070-104033959 | 0.79686944 | 0.91338889 | 0.87168333 | 0.85548889 | 0.69672222 | 0.13434444 | 0.85083333 | 0.74 | 0.79401667 | 0.88 |
| chr10:112836088-112836208 | 0.856 | 0.8838 | 0.9362 | 0.9796 | 0.13052 | 0.505 | 0.9642 | 0.92 | 0.8582 | 0.96 |
| chr10:121243284-121243404 | 0.8705 | 0.867 | 0.8951 | 0.9336 | 0.953 | 0.9456 | 0.8882 | 0.8678 | 0.24352 | 0.94 |
| chr10:122290829-122291145 | 0.947 | 0.80671429 | 0.84671429 | 0.87357143 | 0.89214286 | 0.92571429 | 0.73971429 | 0.91071429 | 0.15228571 | 0.9 |

| | | | | | | | | | | |
|---|---|---|---|---|---|---|---|---|---|---|
| chr10:122468530-122469570 | 0.68453929 | 0.89528571 | 0.80153571 | 0.872 | 0.20912857 | 0.4649 | 0.85485714 | 0.83 | 0.62816429 | 0.92 |
| chr10:124601225-124601663 | 0.90121739 | 0.77204348 | 0.87360217 | 0.66326522 | 0.8573913 | 0.87493478 | 0.04893478 | 0.92554545 | 0.92321739 | 0.92 |
| chr10:132690818-132692045 | 0.95569722 | 0.96037234 | 0.96255319 | 0.96403158 | 0.97366327 | 0.97915464 | 0.96053125 | 0.96396842 | 0.9623913 | 0 |
| chr10:133293143-133293290 | 0.93325 | 0.945 | 0.94875 | 0.95216667 | 0.16548333 | 0.73316667 | 0.86216667 | 0.87 | 0.65783333 | 0.98 |
| chr11:2393788-2394198 | 0.87920588 | 0.93794118 | 0.94182353 | 0.92241176 | 0.95317647 | 0.94717647 | 0.16015882 | 0.86952941 | 0.90941176 | 0.95 |
| chr11:3037172-3037292 | 0.93507143 | 0.93328571 | 0.9465 | 0.82885714 | 0.23304286 | 0.671 | 0.91442857 | 0.89342857 | 0.844 | 0.95 |
| chr11:17828246-17828466 | 0.083925 | 0.87783333 | 0.95733333 | 0.964 | 0.96066667 | 0.97466667 | 0.96616667 | 0.89366667 | 0.85866667 | 0.98 |
| chr11:33286680-33286870 | 0.96235714 | 0.881 | 0.96435714 | 0.95942857 | 0.885 | 0.18781429 | 0.964 | 0.96314286 | 0.90514286 | 0.95 |
| chr11:60182396-60183273 | 0.67130909 | 0.65410909 | 0.66170455 | 0.68196364 | 0.09003636 | 0.40225455 | 0.62076364 | 0.59433636 | 0.59579 | 0.88 |
| chr11:61101960-61102752 | 0.89652778 | 0.9095 | 0.13665556 | 0.62133333 | 0.59966667 | 0.5847 | 0.63558889 | 0.78194444 | 0.8365 | 0.95 |
| chr11:64101632-64101752 | 0.93665 | 0.07897778 | 0.95366667 | 0.97355556 | 0.9572 | 0.9445 | 0.9562 | 0.9373 | 0.921 | 0.94 |
| chr11:65815289-65815449 | 0.8967 | 0.9486 | 0.93564444 | 0.9555 | 0.95588889 | 0.9666 | 0.9478 | 0.84 | 0.16226 | 0.94 |
| chr11:68927954-68928174 | 0.7711 | 0.9344 | 0.951 | 0.953 | 0.15966 | 0.4336 | 0.9498 | 0.76 | 0.9174 | 0.92 |
| chr11:70169503-70170001 | 0.90081818 | 0.90045455 | 0.83 | 0.82218182 | 0.79 | 0.78054545 | 0.84936364 | 0.1623 | 0.83281818 | 0.96 |
| chr11:73405124-73405338 | 0.9211 | 0.9346 | 0.9492 | 0.973 | 0.841 | 0.02488 | 0.92 | 0.94 | 0.946 | 0.96 |
| chr11:75266078-75266198 | 0.73025 | 0.897875 | 0.881625 | 0.878125 | 0.0370625 | 0.62025 | 0.41725 | 0.872875 | 0.849 | 0.93 |
| chr11:75441691-75442165 | 0.88466667 | 0.81541667 | 0.88804167 | 0.90875 | 0.95383333 | 0.93691667 | 0.91216667 | 0.21886667 | 0.74716667 | 0.92 |

| | | | | | | | | | | |
|---|---|---|---|---|---|---|---|---|---|---|
| chr11:76578584-76578766 | 0.04648 | 0.8088 | 0.9408 | 0.9258 | 0.9474 | 0.951 | 0.9376 | 0.82 | 0.861 | 0.96 |
| chr11:118341593-118341872 | 0.9123 | 0.9324 | 0.029405 | 0.8994 | 0.9576 | 0.9488 | 0.931 | 0.95675 | 0.831 | 0.95 |
| chr11:118342540-118343619 | 0.75407143 | 0.83957143 | 0.06562381 | 0.84757143 | 0.866 | 0.879 | 0.76771429 | 0.84180952 | 0.8607619 | 0.89 |
| chr11:118344006-118344398 | 0.6998 | 0.8028 | 0.0358 | 0.9414 | 0.9442 | 0.9412 | 0.917 | 0.8372 | 0.8406 | 0.94 |
| chr11:119024684-119024904 | 0.0553875 | 0.815 | 0.8594375 | 0.861625 | 0.849375 | 0.836625 | 0.9035 | 0.81 | 0.78075 | 0.93 |
| chr11:121570054-121570272 | 0.93464286 | 0.93371429 | 0.91357143 | 0.91314286 | 0.75557143 | 0.94185714 | 0.04451429 | 0.97785714 | 0.93485714 | 0.96 |
| chr12:3791539-3791687 | 0.9403 | 0.9396 | 0.966375 | 0.976 | 0.9582 | 0.9386 | 0.96675 | 0.08 | 0.9034 | 0.86 |
| chr12:4324567-4324841 | 0.96381703 | 0.96066667 | 0.9622971 | 0.96475 | 0.97217391 | 0.964 | 0.95604348 | 0.97 | 0.96791667 | 0 |
| chr12:64613932-64614052 | 0.9297 | 0.9402 | 0.9041 | 0.929 | 0.9188 | 0.9274 | 0.0682 | 0.9544 | 0.9406 | 0.91 |
| chr12:101738826-101739661 | 0.9582543 | 0.95875 | 0.96462171 | 0.96791803 | 0.97198333 | 0.97083051 | 0.96279661 | 0.96 | 0.96452542 | 0 |
| chr12:119699366-119699827 | 0.94936364 | 0.05716364 | 0.9595 | 0.97054545 | 0.96172727 | 0.96427273 | 0.95018182 | 0.89972727 | 0.95609091 | 0.95 |
| chr12:122235766-122235886 | 0.9425 | 0.9424 | 0.9589 | 0.959 | 0.5698 | 0.03228 | 0.9782 | 0.9594 | 0.9408 | 0.93 |
| chr12:122852416-122852907 | 0.73813636 | 0.86036364 | 0.881 | 0.92454545 | 0.94981818 | 0.92481818 | 0.91236364 | 0.16 | 0.88163636 | 0.92 |
| chr12:124424000-124424439 | 0.80777778 | 0.87166667 | 0.93105556 | 0.09523333 | 0.86294444 | 0.63044444 | 0.92566667 | 0.89 | 0.92583333 | 0.96 |
| chr12:124721316-124721436 | 0.6999 | 0.8792 | 0.9463 | 0.9276 | 0.03424 | 0.3278 | 0.9222 | 0.82 | 0.8454 | 0.95 |
| chr12:124874597-124874787 | 0.88904167 | 0.927 | 0.86236742 | 0.85775 | 0.09377273 | 0.39643333 | 0.83125 | 0.87 | 0.86408333 | 0.92 |
| chr12:132115585-132115705 | 0.9379 | 0.9566 | 0.7976 | 0.1744 | 0.961 | 0.941 | 0.9414 | 0.912 | 0.96 | 0.93 |

| | | | | | | | | | | |
|---|---|---|---|---|---|---|---|---|---|---|
| chr12:132288532-132289182 | 0.86994028 | 0.04165833 | 0.90029167 | 0.92275 | 0.93233333 | 0.92436111 | 0.90347222 | 0.85099444 | 0.86638889 | 0.95 |
| chr12:132664891-132665011 | 0.8802 | 0.9418 | 0.9254 | 0.7992 | 0.525 | 0.03734 | 0.977 | 0.9226 | 0.945 | 0.93 |
| chr12:132681195-132681315 | 0.95828571 | 0.95385714 | 0.95464286 | 0.95371429 | 0.85671429 | 0.191 | 0.947 | 0.95742857 | 0.96271429 | 0.94 |
| chr13:45194637-45194857 | 0.81785714 | 0.06207143 | 0.95635714 | 0.96785714 | 0.89971429 | 0.94228571 | 0.94157143 | 0.93 | 0.89042857 | 0.97 |
| chr13:98428885-98429005 | 0.9514 | 0.968 | 0.9454 | 0.9538 | 0.945 | 0.0623 | 0.9168 | 0.95 | 0.903 | 0.93 |
| chr13:110719056-110719904 | 0.94940845 | 0.95728169 | 0.9582089 | 0.95792958 | 0.9696 | 0.9656338 | 0.96718841 | 0.96224286 | 0.95917143 | 0 |
| chr13:111345015-111345235 | 0.88585 | 0.785 | 0.8986 | 0.8853 | 0.9036 | 0.8926 | 0.8974 | 0.2256 | 0.7887 | 0.9 |
| chr13:112559196-112559316 | 0.09823 | 0.886 | 0.9448 | 0.9316 | 0.9476 | 0.865 | 0.8958 | 0.942 | 0.92 | 0.96 |
| chr13:112823434-112823626 | 0.96258929 | 0.1190875 | 0.9610625 | 0.97275 | 0.972375 | 0.97325 | 0.963375 | 0.96 | 0.9495 | 0.96 |
| chr13:113609362-113609482 | 0.85783333 | 0.9185 | 0.94075 | 0.95266667 | 0.82383333 | 0.13916667 | 0.92183333 | 0.87733333 | 0.91233333 | 0.99 |
| chr14:22529329-22529449 | 0.9587 | 0.9702 | 0.07753 | 0.9594 | 0.9812 | 0.9496 | 0.961 | 0.946 | 0.9574 | 0.98 |
| chr14:93067048-93067321 | 0.9391 | 0.04454 | 0.9531 | 0.9552 | 0.9044 | 0.938 | 0.9554 | 0.85 | 0.9088 | 0.94 |
| chr14:98227402-98227522 | 0.6309 | 0.714 | 0.05333 | 0.579 | 0.8668 | 0.8818 | 0.742 | 0.891 | 0.865 | 0.95 |
| chr14:99213304-99213424 | 0.8662 | 0.7112 | 0.14281 | 0.8734 | 0.9206 | 0.9348 | 0.8408 | 0.9196 | 0.8606 | 0.93 |
| chr14:99240664-99240784 | 0.86834524 | 0.93957143 | 0.06501429 | 0.88357143 | 0.91157143 | 0.91857143 | 0.89214286 | 0.93585714 | 0.93383333 | 0.95 |
| chr14:99259433-99259553 | 0.84294444 | 0.89588889 | 0.03498333 | 0.89344444 | 0.91355556 | 0.92266667 | 0.93666667 | 0.93266667 | 0.959 | 0.95 |
| chr14:100689202-100689322 | 0.93375 | 0.83116667 | 0.91291667 | 0.94716667 | 0.928 | 0.93183333 | 0.945 | 0.94 | 0.0709 | 0.97 |

| | | | | | | | | | | |
|---|---|---|---|---|---|---|---|---|---|---|
| chr14:102210488-102211003 | 0.9057962 | 0.93141667 | 0.95014583 | 0.94895833 | 0.83375 | 0.05145 | 0.91045833 | 0.93079167 | 0.95454167 | 0.78 |
| chr14:103581307-103581533 | 0.9139 | 0.8865 | 0.9545 | 0.9632 | 0.9714 | 0.9541 | 0.961 | 0.88 | 0.08507 | 0.94 |
| chr14:104680740-104680860 | 0.75621429 | 0.79442857 | 0.93457143 | 0.928 | 0.77642857 | 0.87314286 | 0.916 | 0.74757143 | 0.10597143 | 0.97 |
| chr14:104922773-104922893 | 0.8457 | 0.8064 | 0.8903 | 0.9462 | 0.9856 | 0.9576 | 0.11326 | 0.8802 | 0.854 | 0.95 |
| chr14:105691549-105691669 | 0.8761875 | 0.88575 | 0.8925 | 0.89975 | 0.93175 | 0.926125 | 0.156325 | 0.884375 | 0.821125 | 0.93 |
| chr14:106200673-106200793 | 0.86003571 | 0.85114286 | 0.05923571 | 0.823 | 0.91842857 | 0.92914286 | 0.88071429 | 0.86 | 0.88614286 | 0.94 |
| chr15:31063054-31063274 | 0.07416667 | 0.796 | 0.834 | 0.9062 | 0.939 | 0.92966667 | 0.90433333 | 0.90193333 | 0.84073333 | 0.95 |
| chr15:40982809-40983029 | 0.10094091 | 0.92063636 | 0.94413636 | 0.92581818 | 0.94063636 | 0.90718182 | 0.93754545 | 0.9 | 0.94109091 | 0.91 |
| chr15:78003549-78003919 | 0.86860556 | 0.804 | 0.95008889 | 0.9609 | 0.9538 | 0.9239 | 0.96255556 | 0.12 | 0.6684 | 0.94 |
| chr15:93413749-93413884 | 0.90791667 | 0.87316667 | 0.87433333 | 0.9075 | 0.863 | 0.912 | 0.86983333 | 0.79 | 0.09465 | 0.96 |
| chr15:101140071-101140191 | 0.9104 | 0.8748 | 0.9046 | 0.8524 | 0.016 | 0.9704 | 0.5156 | 0.9262 | 0.9014 | 0.94 |
| chr16:530129-530349 | 0.90586364 | 0.79836364 | 0.91731818 | 0.17455455 | 0.95172727 | 0.95954545 | 0.94636364 | 0.93 | 0.93427273 | 0.92 |
| chr16:1512265-1512457 | 0.13248 | 0.8518 | 0.91395 | 0.8864 | 0.9119 | 0.9018 | 0.9019 | 0.89 | 0.8896 | 0.96 |
| chr16:1537198-1537418 | 0.07408056 | 0.92961111 | 0.88320425 | 0.92158824 | 0.9025 | 0.89135294 | 0.89127778 | 0.86172222 | 0.88444444 | 0.94 |
| chr16:1550234-1550413 | 0.06132 | 0.8868 | 0.8577 | 0.8934 | 0.9042 | 0.9342 | 0.8658 | 0.8312 | 0.893 | 0.9 |
| chr16:3067320-3068134 | 0.85416667 | 0.76083333 | 0.13638889 | 0.63460556 | 0.93333333 | 0.93522222 | 0.78494444 | 0.82166667 | 0.77316667 | 0.94 |
| chr16:11229880-11230045 | 0.9595 | 0.9508 | 0.9612 | 0.9726 | 0.974 | 0.9618 | 0.9496 | 0.2 | 0.9018 | 0.96 |
| chr16:11682408-11682528 | 0.88635714 | 0.81042857 | 0.94628571 | 0.94557143 | 0.87585714 | 0.91771429 | 0.03155714 | 0.95242857 | 0.92228571 | 0.96 |

| | | | | | | | | | | |
|---|---|---|---|---|---|---|---|---|---|---|
| chr16:14307861-14307981 | 0.76171429 | 0.82585714 | 0.89664286 | 0.91 | 0.93085714 | 0.93114286 | 0.93457143 | 0.79 | 0.11485714 | 0.95 |
| chr16:28986549-28986669 | 0.96207143 | 0.96242857 | 0.0393 | 0.62557143 | 0.95142857 | 0.969 | 0.97428571 | 0.96 | 0.96157143 | 0.96 |
| chr16:81546785-81547005 | 0.9299 | 0.13118 | 0.9591 | 0.9577 | 0.9251 | 0.9251 | 0.9299 | 0.9308 | 0.922 | 0.96 |
| chr16:85642641-85643297 | 0.9238 | 0.74311429 | 0.90427143 | 0.24818286 | 0.76104857 | 0.66102571 | 0.90437143 | 0.91 | 0.90977143 | 0.94 |
| chr16:88653861-88654081 | 0.8350625 | 0.861 | 0.8735 | 0.0864375 | 0.94125 | 0.945 | 0.90675 | 0.82 | 0.854125 | 0.91 |
| chr16:88840759-88840979 | 0.92575 | 0.92755556 | 0.92156944 | 0.96244444 | 0.70188889 | 0.0508625 | 0.91933333 | 0.95833333 | 0.92566667 | 0.93 |
| chr16:88844409-88844543 | 0.8388 | 0.8582 | 0.927 | 0.9544 | 0.9444 | 0.9204 | 0.9104 | 0.2366 | 0.895 | 0.95 |
| chr17:2071895-2072048 | 0.03336364 | 0.92827273 | 0.95809091 | 0.97472727 | 0.91936364 | 0.90136364 | 0.96318182 | 0.81 | 0.89172727 | 0.96 |
| chr17:16860665-16860785 | 0.9401 | 0.8714 | 0.7677 | 0.87 | 0.8716 | 0.7476 | 0.8574 | 0.81 | 0.05828 | 0.96 |
| chr17:18671599-18672260 | 0.96477273 | 0.96312121 | 0.96336364 | 0.96560606 | 0.97093939 | 0.96224242 | 0.96048485 | 0.96272727 | 0.96772727 | 0 |
| chr17:27878804-27879024 | 0.90735714 | 0.882 | 0.13195714 | 0.85628571 | 0.94857143 | 0.92328571 | 0.88742857 | 0.85 | 0.75671429 | 0.94 |
| chr17:29609056-29609276 | 0.89329167 | 0.93483333 | 0.87229167 | 0.19504167 | 0.88916667 | 0.91766667 | 0.93318182 | 0.9 | 0.91866667 | 0.93 |
| chr17:41501981-41503741 | 0.83567778 | 0.90368478 | 0.81880229 | 0.87171111 | 0.90908889 | 0.90847333 | 0.82954 | 0.79 | 0.1228087 | 0.92 |
| chr17:46171194-46171314 | 0.9605 | 0.9478 | 0.9665 | 0.9722 | 0.9264 | 0.0152 | 0.9438 | 0.92 | 0.9348 | 0.95 |
| chr17:63499293-63499413 | 0.6601 | 0.4692 | 0.8673 | 0.8906 | 0.0827 | 0.8178 | 0.7908 | 0.8646 | 0.7512 | 0.94 |
| chr17:74623601-74624169 | 0.74261111 | 0.80088889 | 0.58912778 | 0.60811111 | 0.2171 | 0.54671111 | 0.61828889 | 0.80855556 | 0.76311111 | 0.95 |
| chr17:75543172-75543392 | 0.923 | 0.07153333 | 0.91483333 | 0.93783333 | 0.96616667 | 0.94966667 | 0.9225 | 0.92 | 0.947 | 0.93 |

| | | | | | | | | | | |
|---|---|---|---|---|---|---|---|---|---|---|
| chr17:76328752-76328957 | 0.93059091 | 0.95809091 | 0.96563636 | 0.96081818 | 0.91645455 | 0.18032727 | 0.96909091 | 0.94409091 | 0.95 | 0.94 |
| chr17:76912508-76912728 | 0.9134 | 0.05872727 | 0.92895455 | 0.93672727 | 0.95063636 | 0.94409091 | 0.94845455 | 0.86836364 | 0.88509091 | 0.95 |
| chr17:81192786-81192959 | 0.92503846 | 0.82084615 | 0.94061538 | 0.94161538 | 0.94715385 | 0.93138462 | 0.90730769 | 0.85184615 | 0.20238462 | 0.96 |
| chr17:81270248-81270468 | 0.93627273 | 0.91081818 | 0.90604545 | 0.9056 | 0.7773 | 0.22047273 | 0.92354545 | 0.957 | 0.95736364 | 0.95 |
| chr17:82126627-82127503 | 0.88910884 | 0.89641463 | 0.10988171 | 0.58548537 | 0.93044737 | 0.9185 | 0.84734146 | 0.91 | 0.936925 | 0.88 |
| chr17:82846052-82846272 | 0.22047348 | 0.92158333 | 0.95620833 | 0.96191667 | 0.94741667 | 0.97133333 | 0.96575 | 0.9245 | 0.93808333 | 0.95 |
| chr18:9859284-9859404 | 0.94266667 | 0.93033333 | 0.73283333 | 0.09041667 | 0.901 | 0.8735 | 0.71983333 | 0.92066667 | 0.92033333 | 0.96 |
| chr19:1077845-1077965 | 0.90444444 | 0.77755556 | 0.09106667 | 0.87277778 | 0.69822222 | 0.88722222 | 0.75022222 | 0.92 | 0.92422222 | 0.92 |
| chr19:1151960-1152360 | 0.0524029 | 0.90565217 | 0.95322917 | 0.96170833 | 0.948125 | 0.95166667 | 0.936875 | 0.90566667 | 0.91666667 | 0.94 |
| chr19:1304515-1304635 | 0.88021429 | 0.92985714 | 0.89757143 | 0.93057143 | 0.95185714 | 0.953 | 0.91966667 | 0.815 | 0.13836667 | 0.96 |
| chr19:1370935-1371373 | 0.90790625 | 0.08303438 | 0.95057813 | 0.95732258 | 0.89853125 | 0.9064375 | 0.9314375 | 0.91765625 | 0.93675 | 0.94 |
| chr19:2324602-2324722 | 0.93488889 | 0.91811111 | 0.9409375 | 0.89 | 0.86388889 | 0.81522222 | 0.15073333 | 0.94555556 | 0.951625 | 0.96 |
| chr19:4449605-4449725 | 0.9409 | 0.9372 | 0.9625 | 0.943 | 0.9578 | 0.9778 | 0.972 | 0.19 | 0.9298 | 0.97 |
| chr19:5250020-5250645 | 0.83536957 | 0.82765217 | 0.79132609 | 0.85836087 | 0.85017391 | 0.88592609 | 0.85417391 | 0.17526522 | 0.84430435 | 0.92 |
| chr19:10764202-10764410 | 0.8355 | 0.03578 | 0.9405 | 0.852 | 0.9228 | 0.751 | 0.9026 | 0.807 | 0.8132 | 0.98 |
| chr19:11438041-11438161 | 0.9142 | 0.9424 | 0.9181 | 0.9434 | 0.9154 | 0.935 | 0.0622 | 0.92 | 0.9468 | 0.95 |
| chr19:13002626-13003006 | 0.8976 | 0.8796 | 0.85383333 | 0.87326667 | 0.93706667 | 0.922 | 0.84906667 | 0.09372 | 0.91466667 | 0.96 |
| chr19:13003078-13003434 | 0.89164583 | 0.917375 | 0.89871875 | 0.9455 | 0.9488125 | 0.93525 | 0.9068125 | 0.2 | 0.8285625 | 0.95 |
| chr19:18149454-18149819 | 0.94687 | 0.94876 | 0.94352 | 0.19835417 | 0.88232 | 0.83828 | 0.94812 | 0.92 | 0.92848 | 0.94 |

| | | | | | | | | | | |
|---|---|---|---|---|---|---|---|---|---|---|
| chr19:48967528-48967884 | 0.84783333 | 0.88666667 | 0.94733333 | 0.9375 | 0.0669 | 0.56633333 | 0.72266667 | 0.85416667 | 0.898 | 0.88 |
| chr19:54661889-54662201 | 0.8338 | 0.863 | 0.9061 | 0.9557 | 0.9073 | 0.9613 | 0.17338 | 0.9017 | 0.9051 | 0.86 |
| chr19:55615155-55615865 | 0.95528113 | 0.96142857 | 0.96369333 | 0.97207792 | 0.96418919 | 0.96532895 | 0.96548684 | 0.96272 | 0.95825333 | 0 |
| chr19:55615929-55616819 | 0.95268737 | 0.96076667 | 0.96864254 | 0.97391209 | 0.95916854 | 0.96319101 | 0.96581111 | 0.95686517 | 0.95705682 | 0 |
| chr19:55626556-55626676 | 0.90066667 | 0.921 | 0.04973333 | 0.73183333 | 0.955 | 0.93283333 | 0.71566667 | 0.93566667 | 0.9185 | 0.95 |
| chr20:36645422-36645688 | 0.896 | 0.94442857 | 0.12257143 | 0.59957143 | 0.95542857 | 0.92714286 | 0.95 | 0.92428571 | 0.90057143 | 0.93 |
| chr20:47771130-47771250 | 0.7616 | 0.6372 | 0.7954 | 0.6496 | 0.2246 | 0.9474 | 0.7376 | 0.79 | 0.635 | 0.88 |
| chr20:62186972-62187092 | 0.8729 | 0.9242 | 0.9348 | 0.9442 | 0.8866 | 0.19888 | 0.9488 | 0.8586 | 0.8904 | 0.96 |
| chr20:64001707-64001827 | 0.91941667 | 0.90866667 | 0.96008333 | 0.97 | 0.94816667 | 0.9516 | 0.96516667 | 0.17 | 0.85083333 | 0.96 |
| chr21:37236192-37236312 | 0.937 | 0.96616667 | 0.95433333 | 0.1775 | 0.94666667 | 0.93083333 | 0.95416667 | 0.93383333 | 0.89066667 | 0.97 |
| chr21:37863989-37864109 | 0.9302 | 0.9612 | 0.5273 | 0.7904 | 0.1041 | 0.4448 | 0.6302 | 0.9496 | 0.951 | 0.96 |
| chr21:42660858-42660978 | 0.9691 | 0.7924 | 0.8792 | 0.918 | 0.9644 | 0.9202 | 0.09166 | 0.9374 | 0.9334 | 0.93 |
| chr21:43003359-43003479 | 0.9592 | 0.9524 | 0.9684 | 0.9766 | 0.9822 | 0.946 | 0.9584 | 0.97 | 0.16908 | 0.96 |
| chr21:45348122-45348286 | 0.07145357 | 0.85038462 | 0.86532692 | 0.92207143 | 0.88830769 | 0.91971429 | 0.84121429 | 0.78607143 | 0.77285714 | 0.96 |
| chr21:45531327-45531987 | 0.96225783 | 0.9551 | 0.96226407 | 0.96201724 | 0.97208929 | 0.96561017 | 0.96415789 | 0.967 | 0.94770909 | 0 |
| chr22:21689890-21690010 | 0.83583333 | 0.83783333 | 0.83641667 | 0.08586667 | 0.852 | 0.855 | 0.86316667 | 0.855 | 0.707 | 0.94 |
| chr22:23265333-23265529 | 0.90925 | 0.12733333 | 0.96166667 | 0.955 | 0.96983333 | 0.96366667 | 0.963 | 0.97116667 | 0.93833333 | 0.93 |

| | | | | | | | | | | |
|---|---|---|---|---|---|---|---|---|---|---|
| chr22:37144234-37144469 | 0.7935 | 0.91316667 | 0.85725 | 0.16913333 | 0.8795 | 0.87333333 | 0.8195 | 0.68366667 | 0.75 | 0.95 |
| chr22:45996289-45996409 | 0.6037 | 0.4326 | 0.8408 | 0.8394 | 0.11322 | 0.5242 | 0.4376 | 0.91 | 0.8914 | 0.97 |
| chr22:46214216-46215152 | 0.95734426 | 0.95653968 | 0.96325642 | 0.96957627 | 0.97001724 | 0.96996721 | 0.96089831 | 0.97 | 0.96253333 | 0 |
| chr22:46416434-46416554 | 0.9423 | 0.07752 | 0.9073 | 0.9714 | 0.9814 | 0.9748 | 0.9586 | 0.9188 | 0.92 | 0.98 |
| chr22:48079853-48079973 | 0.85416667 | 0.69933333 | 0.1199 | 0.60866667 | 0.90083333 | 0.90183333 | 0.81216667 | 0.84333333 | 0.84266667 | 0.96 |
| chrX:150870885-150871005 | 0.02708 | 0.846 | 0.8759 | 0.8606 | 0.855 | 0.7966 | 0.8228 | 0.92 | 0.9 | 0.93 |
